# OH-EpiCap: a semi-quantitative tool for the evaluation of One Health epidemiological surveillance capacities and capabilities

**DOI:** 10.1101/2023.01.04.23284159

**Authors:** Henok Ayalew Tegegne, Carlijn Bogaardt, Lucie Collineau, Géraldine Cazeau, Renaud Lailler, Johana Reinhardt, Emma L. Taylor, Joaquin Prada, Viviane Hénaux

## Abstract

Although international health agencies encourage the development of One Health (OH) surveillance, many systems remain mostly compartmentalized, with limited collaborations among sectors and disciplines. In the framework of the OH European Joint Programme “MATRIX” project, a generic evaluation tool called OH-EpiCap has been developed to enable individual institutes/governments to characterize, assess and monitor their own OH epidemiological surveillance capacities and capabilities. The tool is organized around three dimensions: organization, operational activities, and impact of the OH surveillance system; each dimension is then divided into four targets, each including four indicators. A semi-quantitative questionnaire enables the scoring of each indicator, with four levels according to the degree of satisfaction in the studied OH surveillance system. The evaluation is conducted by a panel of surveillance representatives (during a half-day workshop or with a back-and-forth process to reach a consensus). An R Shiny-based web application facilitates implementation of the evaluation and visualization of the results, and includes a benchmarking option. The tool was piloted on several foodborne hazards (i.e. *Salmonella, Campylobacter, Listeria*), emerging threats (e.g. antimicrobial resistance) and other zoonotic hazards (psittacosis) in multiple European countries in 2022. These case studies showed that the OH-EpiCap tool supports the tracing of strengths and weaknesses in epidemiological capacities and the identification of concrete and direct actions to improve collaborative activities at all steps of surveillance. It appears complementary to the existing EU-LabCap tool, designed to assess the capacity and capability of European microbiology laboratories. In addition, it provides opportunity to reinforce trust between surveillance stakeholders from across the system and to build a good foundation for a professional network for further collaboration.

## 1 Introduction

In recent years, the One Health (OH) concept has gained momentum, and international efforts have been made to strengthen the implementation of multi-sectoral surveillance to more effectively manage health hazards at the human, animal and environment interface (1). OH surveillance is a collaborative and systematic collection, validation, analysis, interpretation of data, and dissemination of information collected on humans, animals, and the environment from different sectors and disciplines to inform decisions for more effective evidence-based health interventions (2,3). However, in spite of the efforts of the quadripartite alliance between the Food and Agriculture Organisation of the United Nations (FAO), the World Organisation for Animal Health (WOAH-OIE), the World Health Organisation (WHO), and the United NationsEnvironment Programme (UNEP) (4) to promote collaboration in surveillance and laboratory networks and overpass professional silos, most surveillance systems remain compartmentalized, with limited interaction across actors in the system (5). For multiple reasons, implementing OH approaches in practice still proves challenging (6) and collaborations between health sectors occur mostly in crisis times (7).

There is a wide range of possible organizational models for collaboration, and its operationalization varies in terms of areas of implementation throughout the surveillance process (8–11). Collaboration is mainly driven by the epidemiological context and surveillance objective and is built according to actors’ expectations (5). Regular evaluation of the organization and functionality of collaboration is crucial to assess the surveillance system’s capacity and capability to produce relevant information, identify areas for improvement, and optimize added value gained by integrating efforts across sectors.

In recent years, several methods have been developed to assess whether collaborative efforts are appropriate and functional and whether it improves the impact of surveillance systems (12,13). The Evaluation of Collaboration for Surveillance (ECoSur) tool targets the organization and functioning of multi-sectoral collaborations in a surveillance system (5). It relies on a semi-quantitative approach, with data collection based on interviews of the coordinators of the programs included in the surveillance system, requiring a one-to-two-week evaluation period on average (5). The Network for Evaluation of One Health (NEOH) relies on the theory of change to identify the necessary preconditions and actions to be taken to reach long-term goals (14). The whole process is estimated to take one to two months and requires interviews of essential actors and stakeholders (13). The OH Assessment for Planning and Performance (OH-APP) focuses on multi-sectoral coordination mechanisms to inform planning and development assistance. The OH-APP complements the WHO Joint External Evaluation by providing specific indicators to measure the maturity of a multi-sectoral coordination mechanism and benchmark its progress toward a sustainable mechanism capable of coordinating multi-sectoral and multi-stakeholder collaboration for preparedness and response to public health threats (https://www.onehealthapp.org/about). Other tools were developed specifically for antimicrobial resistance (AMR) surveillance activities: the Progressive Management Pathway tool for AMR (PMP-AMR), the AMR integrated surveillance system evaluation project (ISSEP) tool, the Assessment Tool for Laboratories and AMR Surveillance Systems (ATLASS) (13) and the Integrated Surveillance System Evaluation (ISSE) framework (2). The different tools appear complementary in terms of evaluation objectives and provide generic science-based guidance for the evaluation of collaboration in surveillance systems. Yet, they also appear quite complex and require a lot of data, time, and human resources (13), limiting their (regular) implementation.

The OH European Joint Programme MATRIX project aims to produce guidelines and tools applicable at the national level to connect existing surveillance structures and resources, and strengthen integrated surveillance initiatives, ultimately adding value by building on existing resources, and creating synergies among sectors. In this context, we developed a generic evaluation and benchmarking tool (OH-EpiCap), implemented through an interactive online web application, for characterizing, monitoring, and evaluating epidemiological national surveillance capacities and capabilities for OH surveillance. This tool was designed to enable representatives of any surveillance system to conduct an evaluation of the multiple aspects of OH surveillance, in a short time and without requiring an external evaluation team. The evaluation addresses the multisectoral and multidisciplinary efforts to ensure communication, collaboration, and coordination among all relevant actors of the surveillance working locally, nationally, and globally to attain optimal health for people, animals, and our environment (https://extranet.who.int/sph/one-health-operations). Besides identifying areas that could lead to improvements in existing OH epidemiological surveillance capacities, the tool was designed to allow benchmarking (i.e. comparisons) with results from previous evaluations of that surveillance system, or other relevant systems, for example in other countries.

## 2 Methods

### 2.1 Identification, definition and validation of indicators

Existing evaluation tools focusing on multi-sectoral and interdisciplinary collaboration aspects in epidemiological surveillance were used as a basis for the development of the OH-EpiCap tool. Besides, to structure our tool, we considered the format of the EU-LabCap tool, developed to assess bi-annually the capacity and capabilities of European microbiology laboratories (15).

Three dimensions of evaluation were considered in our tool: the organization of the collaborative system, the nature and functioning of collaborations for operational activities, and the impact of collaborations on surveillance. Each dimension was then divided into several targets focusing on specific features of multi-sectoral collaborations, building from the existing evaluation frameworks. Finally, we established standardized indicators defining more accurately each target and we singled out the necessary criteria to support their evaluation.

The organization and definition of the targets and indicators were consolidated and validated through expert consultation. Experts were selected based on previous and ongoing involvement in research activities on OH aspects (e.g. One Health - European Joint Project (OH-EJP) program; Convergence in evaluation frameworks for integrated surveillance of AMR (CoEvalAMR) project) in national veterinary and / or public health institutes and from EFSA. The experts were asked to review and comment on all the proposed indicators and identify missing information. The initial list of indicators was refined based on experts’ comments and validated with them through a back and forth process. Additional specific modifications were also carried out based on feedback from participants in case studies during the pilot phase (see below).

### 2.2 Questionnaire and semi-quantitative scoring options

A questionnaire was developed to facilitate the collection of information for the scoring of the indicators, with one question per indicator. A semi-quantitative scale was defined with four levels, describing the level of compliance of the system under examination compared to an ideal situation: higher values suggest better adherence to the OH principle targeted by the indicator (i.e. better integration of sectors) and lower values indicate improvements may be beneficial. In addition, the option of “Not applicable” (NA) was included to take into consideration the case where the indicator would not be relevant to the OH surveillance system under evaluation. The scoring defined for each indicator is available at the end of the user guide (16).

### 2.3 Data visualization and web application

A web application was developed (using R *shiny* and *shinydashboard* packages) (17), (18)with a user guide describing the different steps for completing the questionnaire and visualizing the results (16). The link to the application is: https://freddietafreeth.shinyapps.io/OH-EpiCap/. The interface enables users to complete the questionnaire interactively (and also to upload the answers from a questionnaire completed previously). Below each question, free text space is provided to add notes or justify the answer provided. These comments are saved and can be also visualized when reviewing the results of the evaluation. The application allows the user to save partially completed questionnaires in .csv (human-readable) format, to revisit or complete the answers at a later time. The OH-EpiCap web application does not keep any data, to comply with the European General Data Protection Regulation. Users must save their work locally (i.e. in the machine they are using) before closing the application (to avoid any data loss).

The application facilitates the exploration of the completed (and/or uploaded) assessment and of the results of the evaluation by way of multiple visualizations. The answers to the OH-EpiCap questionnaire are analyzed at the target level for each dimension by averaging the scores across the indicators to get a final score (between 1 and 4), and at the dimension level by averaging target-level outputs (the mean scores over all questions are expressed as a percentage). Results are displayed in the form of interactive radar charts and lollipop plots to identify strengths and weaknesses at both dimension and target levels. Users may hover over data points to explore the breakdown of scores for each target and indicator. At the target level, this option displays for each data point the comments provided by the evaluators during the filling of the related question. Finally, users can download a two-page report (in Html format) comprising the graphic outputs and comments highlighting the main strengths and weaknesses of the surveillance system examined. Moreover, the tool also includes a benchmarking functionality to compare results from the ongoing evaluation with a reference set based on results from previous OH-EpiCap evaluations. This reference dataset can be generated from other evaluations that the user has access to, using a specific tab of the web application. This function allows the integration of multiple evaluations (for example, from other countries for the same hazard), thus anonymizing the results for a given system/hazard.

### 2.4 Evaluation Process

The OH-EpiCap tool was designed to serve as a support for discussion and scoring of the OH aspects by a panel of representatives from the different sectors across the entire surveillance system of a specific hazard. The selected surveillance representatives form an evaluation panel, which gathers during a four-hour workshop to complete the questionnaire. For each question, the panel must provide one answer after reaching a consensus. In the case where it is not possible to organize a workshop to conduct the evaluation, the questionnaire may be filled sequentially by the surveillance representatives from each sector, with a back-and-forth process to reach a consensus. Once completed, the online application allows the panel to visualize the outcomes in real-time and to generate a OH-ness profile for the studied system.

### 2.5 Pilot Phase

The OH-EpiCap tool was piloted through several applications on surveillance systems of specific hazards targeted by the MATRIX consortium, including foodborne and other emerging zoonotic hazards. As a first step, for each surveillance system, a representative was identified directly within the MATRIX participants or their professional networks. Then, a one-hour meeting with the identified surveillance representatives was organized to present the tool and the evaluation process, and to answer questions. Participants were then asked to identify additional surveillance representatives to include in the evaluation panel. The choice of conducting a workshop or completing the questionnaire sequentially by representatives was left to the participants.

The OH-EpiCap evaluations conducted through a workshop were conducted in the language of the country to facilitate discussions. One or two persons from the MATRIX research team also participated as observers, to identify areas for improvement in the questionnaire and the evaluation process, and to provide additional explanations if needed during the completion of the questionnaire by participants.

At the end of the workshop, participants were asked to share their thoughts on the evaluation process, the relevance of the evaluation, and any feedback and comments to improve the tool. A checklist was provided for collecting this information regarding the questionnaire and its implementation (Supplementary file S1). The checklist was also provided to participants who conducted the evaluation through a sequential completion of the questionnaire; the time dedicated to the evaluation (for completing the questionnaire and analyzing results) was also requested. Based on the participants’ comments, the questionnaire and scoring options were further improved.

### 2.6 Ethical approval

The MATRIX project went to ethical approval by the ethical advisors of the One Health European Joint Programme. We informed verbally and through email the participants about the following points: 1) the use of the OH-EpiCap tool and application is voluntary; 2) the OH-EpiCap tool does not collect personal information, to comply with the European General Data Protection Regulation; 3) the web application does not keep the data regarding the OH surveillance system evaluated, and does not store them outside of the user’s computer.

## 3 Results

### 3.1 OH-EpiCap Structure

The organization of the OH-EpiCap tool is described in Figure 1. The first dimension, is related to the organization of the OH surveillance system: Target 1.1 *Formalization* focuses on the common aim of the system, support documentations, coordination roles, and leadership in the OH surveillance system; Target 1.2 *Coverage* addresses whether the surveillance covers all relevant sectors, disciplines, actors, geography, populations and hazards; Target 1.3 *Resources* addresses aspects related to financial and human resources, sharing of the available operational resources, and training; and Target 1.4 *Evaluation* focuses on internal and external evaluations, implementation of corrective measures, and the capacity of the OH surveillance system to adapt to changes.

**Figure 1.**
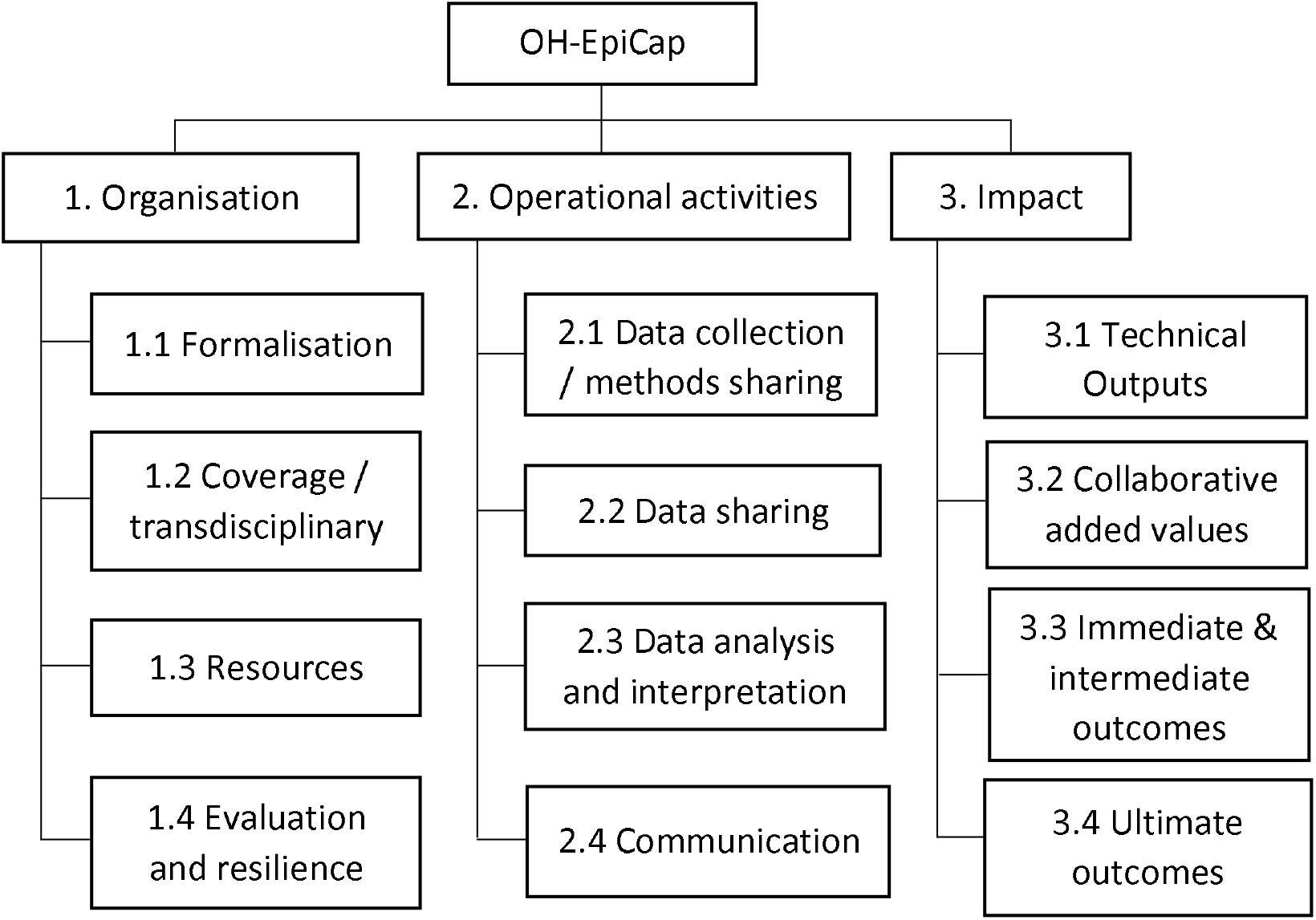
Structural overview of the OH-EpiCap targets, grouped by dimension.

The second dimension deals with OH aspects in operational activities: Target 2.1 D*ata collection and methods sharing* concerns the level of multi-sectoral collaboration in the design of surveillance protocols, data collection, harmonization of laboratory techniques and data warehousing; Target 2.2 *Data sharing* addresses data sharing agreements, evaluation of data quality, use of shared data, and the compliance of data with the FAIR principle; Target 2.3 *Data analysis and interpretation* addresses multi-sectoral integration for data analysis, sharing of statistical analysis techniques, sharing of scientific expertise, and harmonization of indicators; and Target 2.4 *Communication* focuses on both internal and external communication processes, dissemination to decision-makers, and information sharing in case of suspicion.

The third dimension deals with the impact of the OH surveillance system: Target 3.1 *Technical outputs* concerns the timely detection of emergence, knowledge improvement on hazard epidemiological situations, increased effectiveness of surveillance, and reduction of operational costs.; Target 3.2 *Collaborative added value* addresses strengthening of the OH team and network, international collaboration and common strategy (road map) design; Target *3*.*3 Immediate and intermediate outcomes* addresses advocacy, awareness, preparedness and interventions based on the information generated by the OH surveillance system; and Target *3*.*4 Ultimate outcomes* focuses on research opportunities, policy changes, behavioral changes and better health outcomes that are attributed to the OH surveillance system.

The list of indicators in each target and the standardized scoring guide detailing, for each individual score, the situation in which that score should be awarded, is available in Hénaux et al. (16).

### 3.2 Pilot OH-EpiCap applications

Eight evaluations were conducted during the pilot phase of the OH-EpiCap tool; these assessments targeted the multi-sectoral surveillance system for antimicrobial resistance (AMR) in France and in Portugal, *Salmonella* in France, Germany, and the Netherlands, *Listeria* in the Netherlands, *Campylobacter* in Sweden, and psittacosis in Denmark (https://icahs4.org/fileadmin/user_upload/ICAHS4_2020/abstractbook_10maj.pdf, p. 211).

Evaluations of the surveillance systems for psittacosis in Denmark, *Campylobacter* in Sweden and *Salmonella* in Germany were conducted through a half-day workshop. The workshop in Denmark was held in person, and gathered seven surveillance representatives, from the public health sector with expertise in laboratory/bacteriology and epidemiology, and from the animal health sector from the official sampling, laboratory, and risk management unit. The workshop lasted three hours (including a participants and workshop introduction, the filling of the three dimensions of the questionnaire, the results analysis, and debriefing). The workshop in Germany was held online and gathered ten representatives, from the public health (Robert Koch Institute), animal health (Friedrich Loeffler Institute), and food safety (German Federal Institute for Risk Assessment - BfR) sectors. The workshop lasted four hours; the two last targets of the third dimension were not completed during the workshop because of time constraints (and scoring for these indicators was provided at a later stage). The workshop in Sweden was held online and gathered five representatives from the public health (Folkhalsomyndigheten), animal health (National Veterinary Institute - SVA), and food safety (Swedish National Food Agency - SLV; Swedish Board of Agriculture) sectors. The workshop lasted three hours.

Other study cases were conducted through completion of the questionnaire (in a Word format) either by one representative from each sector of surveillance, sequentially (AMR in Portugal and AMR in France), or by one-to-two representatives from one sector only but with a good knowledge of surveillance across sectors and existing multi-sectoral collaborations (*Salmonella* in France, *Listeria* and *Salmonella* in the Netherlands). Then, the OH-EpiCap team recorded the scores in the web application to generate the final report (displaying the results; Supplementary file S2), that was sent to the surveillance representatives. In those study cases, each evaluator spent between two and three hours completing the questionnaire or reviewing and completing a pre-filled questionnaire (in the case of sequential completion).

As observers during the two workshops, we received comments and questions from the panel of the evaluation. Similarly, for the evaluations conducted separately by surveillance representatives, many comments were provided directly in the document (Word questionnaire) to justify the choice of the scores and propose some evolutions of the system for the aspects with a low score. The comments from the panels and suggestions from the MATRIX team are summarized in the table below (Table 1).

**Table 1.** Comments and questions from the surveillance representatives who conducted a OH-EpiCap evaluation and answers from the MATRIX team

| Comments / Questions | Answers |
| --- | --- |
| Answers proposed for a question may not fit the OH surveillance system under evaluation. | The feedback from the evaluators helps refine and complete the answers proposed for some indicators to consider specific OH surveillance contexts and situations not envisaged initially. In addition, the “NA” answer can be used if the question is not relevant for the OH surveillance system under evaluation.<br>If the answers proposed for a question do not fit the OH surveillance system under evaluation, the panel should define what would be the ideal situation and score the question accordingly by comparing the current situation to the ideal one. The panel can specify in the free comment space which alternative answer(s) were considered, for result interpretation and dissemination. |
| Questions about the amount of data that can be saved in the web application and whether the data is accessible by stakeholders not involved in the evaluation, arguing that some information could be potentially confidential. | The web application does not keep any data, to comply with the European General Data Protection Regulation. Users must save their work locally (i.e. in the machine they are using) before closing the application (to avoid any data loss). |
| A question whether the surveillance system under evaluation could be considered as an OH surveillance system in spite of a lack of formalisation or applicable legislation regarding the collaborations between sectors. | The OH-EpiCap tool was developed for any surveillance system where some collaborations between sectors exist (at any step of the surveillance) even if those ones are not formalised or occur occasionally. |
| Integration could be considered from a system-wide perspective, including multi-sectoral collaborations but also inter-program collaborations even within the same sector (e.g. collaborations between a surveillance program targeting AMR and another one on antimicrobial use, in a specific sector) | Although this vision appears different from the OH approach, the tool allows different levels of integration to be considered; however, such specificity should be clearly stated and understood by all surveillance representatives before the start of the evaluation. |
| Need more time to further discuss and map the actions to be taken to improve identified weaknesses | Participants are encouraged to further discuss and investigate underlying issues to improve collaboration in the system, during a dedicated workshop |

## 4 Discussion

The OH-EpiCap tool is a semi-quantitative evaluation tool developed for macro analysis of the OH capacities and capabilities of a system for surveillance of a specific hazard. This tool helps, without a priori consideration, characterize how multi-sectoral collaborations operate within surveillance systems. It facilitates the identification of strengths and weaknesses, focusing on the organization and functioning of existing collaborations, and of their impacts on the effectiveness of surveillance.

The OH-EpiCap tool is generic and can be applied to the surveillance of any hazard. Accordingly, the tool was applied to a large range of hazards, including food-borne hazards (*Salmonella, Listeria* and *Campylobacter*), other zoonotic hazards (psittacosis) and AMR. The questionnaire includes specific indicators oriented towards OH preparedness and response and is therefore of interest for surveillance systems targeting emerging or exotic zoonoses. The pilot phase was beneficial to make the questionnaire more flexible to the diversity of contexts of surveillance, depending on hazards and countries, and to the level of integration of the system. Given that the tool is generic, the importance of clearly specifying the outline of the system under study and the levels of integration considered (e.g. inter-program collaborations), in addition to multi-sectoral integration, is a priority.

Besides, the tool can address any surveillance system, whether they are well formalized or at a low level of integration, as long as some multi-sectoral collaborations exist at any step of the surveillance, even if they are not supported by official regulations, nor formalized through specific agreements and procedures. The formalization of the organization and functioning of the collaborations between sectors is considered an important aspect for OH surveillance (11), and therefore a lack of formalization will lead to low scores in some indicators of the OH-EpiCap tool (in particular in dimension 1). Depending on the aim of the OH surveillance system and if this lack of formalization is considered as an issue, surveillance representatives are encouraged to determine what elements would elevate the current multi-sectoral collaboration level to an official OH surveillance system.

The first step of an OH-EpiCap evaluation process is the identification of the panel of representatives of the surveillance system under study, i.e. who will conduct the evaluation. The composition of the evaluation team must be representative of the whole surveillance system (as much as possible). Thus, the panel should include experts from all sectors involved in the surveillance of the hazard under evaluation, and would encompass a large range of disciplines and experiences regarding the functioning of collaborations among institutes and programs. A mapping of the surveillance system under study, characterizing the programs and institutes involved in the surveillance for each sector, would help identify surveillance representatives. This panel will then work closely together during the evaluation workshop, with ideally all representatives having the opportunity to express their views during the scoring of the indicators. Therefore, identifying respected and well-known members of the surveillance system under study is an asset to moderate respectful discussion and prevent any stronger opinions from monopolizing the exchanges over the quieter contributors.

The second step consists in the evaluation of the OH epidemiological capacities and capabilities following the three dimensions, through the web app. The evaluation is based on a semi-quantitative method; this is certainly marked by subjectivity, especially in the case of a limited panel of evaluators. Indeed, some indicators might be scored very differently across surveillance representatives with various backgrounds, perceptions, and expectations. Yet, we stress that only one answer can be provided to each question; therefore the surveillance representatives of the evaluation panel must reach a consensus to answer each question (based on their backgrounds, perceptions, and expectations). This constraint of having to reach a consensus for each question, within a standardized set of answers, limits the bias of subjectivity. Another limitation of this tool is that the current implementation assumes that all indicators are of equal importance (i.e. have the same weight). This is obviously a simplification and depending on the context of surveillance and the overall aim of the collaborations among sectors, some aspects of the evaluation may appear more important and should therefore get more focus during the result analysis and interpretation, as well as for prioritizing recommendations.

The organization of the evaluation in three distinct parts (one per dimension) helps the panel to articulate its reflection regarding the OH-ness of their surveillance system. It supports a collective and transparent evaluation approach, and facilitates identification of weaknesses and alternatives. Recommendations and concrete actions to improve the global systems can emerge from this process, facilitating in a second step prioritization among actions to improve OH-ness. The user-friendly web app provides a set of classical graphs (gauges, radar charts, lollipop plots) that enables users to easily visualize and analyze the strengths and weaknesses at the level of the indicators, and also of each target within the three dimensions. We underline the importance of taking careful notes during the workshop. Justifications provided by the panel in the comment spaces during questionnaire completion are displayed on the graph, facilitating the interpretation of the results at the end of the evaluation workshop, and also at a later stage as needed (thanks to the options to save and upload results in the web application). A careful documentation of how the questions were interpreted and answered is also recommended to follow changes in the monitoring system over time, through new evaluations by the same panel or by another panel of evaluators.

Securing a half-day window for the workshop would enable the evaluation to be conducted, a report to be generated, and results to be analyzed. However, we stress that further discussions regarding prioritization and planning of actions to improve identified weaknesses, should be scheduled at another time. Based on the evaluations conducted, we observed that the tool provides a manageable “first step for action” where there is an interest in upgrading or renewing existing collaborations across surveillance systems. The OH-EpiCap tool provides a macroscopic analysis of the overall organization, functioning and impact of multi-sectoral collaborations. In some cases, it may be relevant to complement the OH-EpiCap approach with a more thorough evaluation of the weaker OH aspects, using evaluation tools dedicated to the functioning and performance of surveillance (19) and/or OH aspects (13). Besides, the OH-EpiCap tool does not assess OH capacities related to laboratory activities; we recommend to consider applying the OH-LabCap tool (developed within the OH-HARMONY-CAP; https://onehealthejp.eu/jip-oh-harmony-cap/) for such aspects.

Evaluations with the OH-EpiCap tool require little human and time resources; the evaluation can be conducted through a half-day (3-4 hours) workshop, and we recommend limiting the evaluator panel to 8-10 representatives. Evaluations with ECoSur or NEOH require numerous individual interviews with actors of the surveillance, followed by a full-day meeting to validate the results (5,14). A Delphi-like approach (i.e. each representative completes the questionnaire, then a facilitator collates and summarizes all responses, and provides the summary back to the participants for cross-checking/validation) is an alternative option to conduct the evaluation (20). This approach does not enable surveillance representatives to share their views and experiences regarding OH surveillance, in contrast to a roundtable discussion. Therefore such an approach should be preferred in situations where an evaluation would be requested by policymakers within a short delay, for example during surveys assessing the OH epidemiological capacities of EU countries for a specific hazard, or within a country for a large range of related hazards. As such, the tool will be very complementary to the existing EU-LabCap tool, designed to assess the capacity and capability of European microbiology laboratories (15). We emphasize that the benchmarking module of the OH-EpiCap web app enables each country to compare their results to a reference set that could be generated by the policymakers using a compilation of evaluation results for the same hazard from other countries, or for other hazards from the same country, depending on the context.

## 5 Conclusion

OH-EpiCap is a generic (i.e. applicable to multi-sectoral surveillance systems of any hazard), interactive (facilitating and supporting discussions among stakeholders from diverse sectors and disciplines), and standalone (thanks to the user-friendly web application) tool developed to conduct macro-level evaluation of epidemiological national capacities and capabilities for OH surveillance. It supports the diagnostic of strengths and weaknesses in multi-sectoral collaborations and helps to identify concrete and direct actions to improve collaborative activities at all steps of surveillance. Besides, this evaluation framework strengthens trust between stakeholders across the systems, building a foundation for professional networks, acculturation to practices in other health sectors and disciplines, and long-term collaborations.

## Data Availability

All data produced in the present study are available upon reasonable request to the authors

## 6 Acknowledgments

We thank the OH-EJP MATRIX consortium, experts from other OH-EJP projects, the CoEvalAMR project, and the European Food Safety Authority (EFSA) and the participants to the OH-EpiCap evaluations for their feedback and support to consolidate and validate the organization of the tool and the definition of the targets and indicators.

## 7 Funding

This work was supported by funding from the European Union’s Horizon 2020 Research and Innovation programme under grant agreement No 773830: One Health European Joint Programme. Funding sources did not affect the design of this study, data collection, data analysis, decisions on publication, or preparation of the manuscript.

## Notes

### Competing Interest Statement

The authors have declared no competing interest.

### Funding Statement

This study was funded by the European Union Horizon 2020 Research and Innovation programme under grant agreement No 773830: One Health European Joint Programme.

