## Supplementary file S1 for "OH-EpiCap: a semi-quantitative tool for the evaluation of One Health epidemiological surveillance capacities and capabilities"

**Supplementary file S1.** Checklist of points considered by the evaluators during a OH-EpiCap evaluation to provide feedback to the MATRIX team

- Check if the instruction to complete the tool is clear.
- Provide feedback on any potential problem with wording or contents.
- Check that all respondents are interpreting the questions in the same way.
- Check if any question requires additional information (e.g. definitions, examples).
- Check that questions are not leading or biased.
- Check that questions are not too vague (and too long to be answered).
- Check that questions are not redundant.
- Check that the ordering of questions within sections is logical.
- Does all the questions listed are relevant to the evaluation of One Health (multi-sectoral) surveillance capacity?
- Are important aspects regarding the evaluation of One Health (multi-sectoral) surveillance capacities missing?
- Indicate how long it takes to complete the survey.
- If the evaluation was conducted through a dedicated workshop, including surveillance representatives of the system under study, could you please indicate whether any sector, discipline, expertise, etc. appears to be not represented.
- Please provide any additional recommendations or suggestions that can help us improve the EpiCap tool.
