## Supplementary file S2 for "OH-EpiCap: a semi-quantitative tool for the evaluation of One Health epidemiological surveillance capacities and capabilities"

**Supplementary file S2.** Example of a OH-EpiCap report generated by the web application, displaying graphical representations of the evaluation results at the dimension, target and indicator levels.

### OH-EpiCap Index and Dimensions

OH-EpiCap and Dimension indices represent mean scores over all questions, expressed as a percentage.

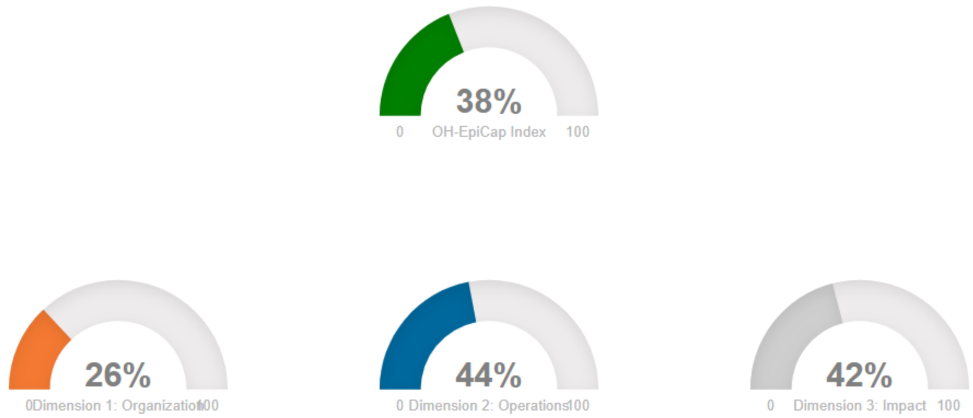


### Targets

This section shows the results of the twelve targets, divided across the three dimensions.
Scores range 1-4, with higher values suggesting better adherence to the One Health principle (better integration of sectors), and lower values suggesting improvements may be beneficial.
Users are encouraged to hover over data points to view a breakdown of each target score.

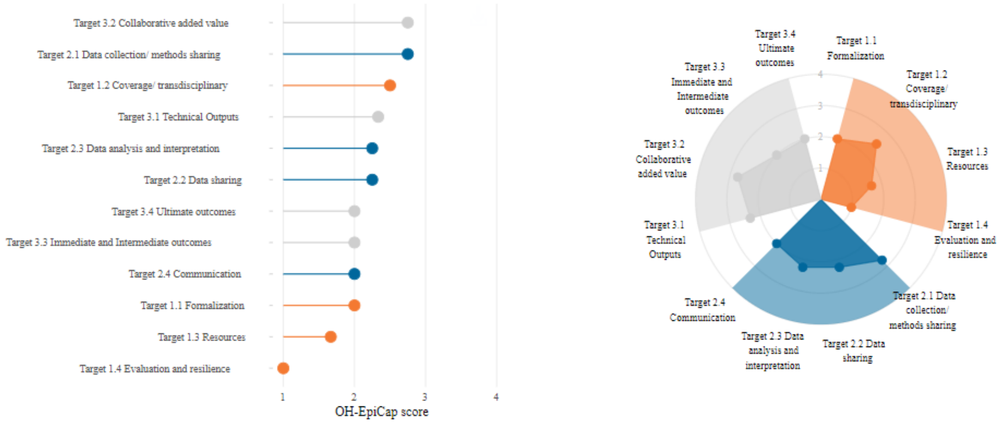


Targets demonstrating **good adherence** to One Health principles are: **None**.
Targets that would **most benefit from improvement** are: **Target 1.3 Resources, Target 1.4 Evaluation and resilience**.

### Dimension 1: Organization

This section shows the results across all indicators within the four targets of Dimension 1 (Organization).

Scores range 1-4, with higher values suggesting better adherence to the One Health principle (better integration of sectors), and lower values suggesting improvements may be beneficial.

Indicators labelled in grey indicate a question was answered with NA. Users are encouraged to hover over plotted data points to view the wording of the chosen indicator level, and any comments that may have been added in connection with a particular question.

Indicators demonstrating **good adherence** to One Health principles are: **None**.

Indicators that would **most benefit from improvement** are: **Supporting documentations, Budget, Training, Internal evaluation, External evaluation, Adaptability to changes**.

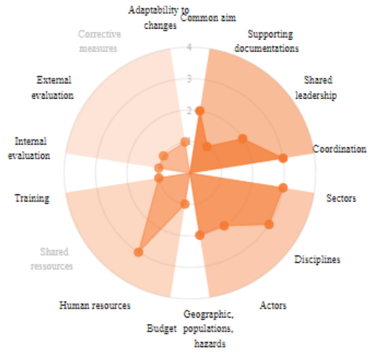


### Dimension 2: Operations

This section shows the results across all indicators within the four targets of Dimension 2 (Operations).

Scores range 1-4, with higher values suggesting better adherence to the One Health principle (better integration of sectors), and lower values suggesting improvements may be beneficial.

Indicators labelled in grey indicate a question was answered with NA. Users are encouraged to hover over plotted data points to view the wording of the chosen indicator level, and any comments that may have been added in connection with a particular question.

Indicators demonstrating **good adherence** to One Health principles are: **Laboratory techniques, External communication**.

Indicators that would **most benefit from improvement** are: **Data collection, Usefulness, Indicators, Internal communication, Emergence**.

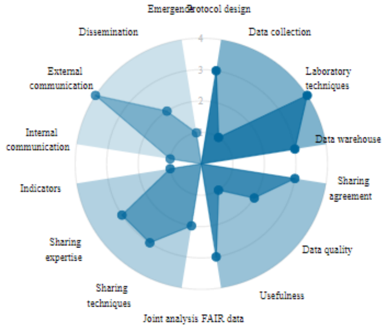

### Dimension 3: Impact

This section shows the results across all indicators within the four targets of Dimension 3 (Impact).

Scores range 1-4, with higher values suggesting better adherence to the One Health principle (better integration of sectors), and lower values suggesting improvements may be beneficial.

Indicators labelled in grey indicate a question was answered with NA. Users are encouraged to hover over plotted data points to view the wording of the chosen indicator level, and any comments that may have been added in connection with a particular question.

Indicators demonstrating **good adherence** to One Health principles are: **Effectiveness, Strategy**.

Indicators that would **most benefit from improvement** are: **Emergence detection, Interventions, Health outcome**.

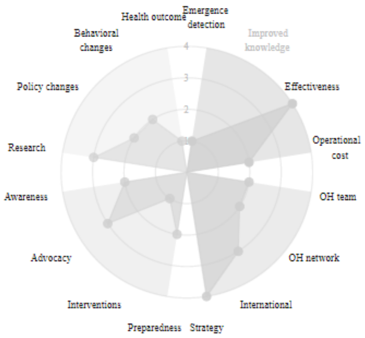
